# Predicting factors of heart arrhythmia in patients with permanent heart pacemaker

**DOI:** 10.1101/2024.11.07.24316937

**Authors:** Thanh Tri Vu, Lac Duy Le, Hoang Minh Vo Nguyen

## Abstract

**Purpose:** Identify some factors predicting arrhythmia after permanent pacemaker placement to treat atrioventricular block and sinus node dysfunction.

**Methods:** Prospective study in 312 patients with permanent pacemakers due to atrioventricular block and sinus node dysfunction being monitored at Thong Nhat Hospital and Thu Duc city hospital from March 2022 to September 2022

**Results:** The rate of arrhythmia after pacemaker insertion was 41%, of which 20.2% atrial fibrillation; 10.9% atrial tachycardia; 1.3% sinus arrest; 9.9% ventricular premature beats; 1.3% ventricular tachycardia and 2.2% supraventricular tachycardia. Factors that determine the possibility of arrhythmia after permanent pacemaker placement include ejection fraction index, blood potassium, blood chloride, blood HDL-Cholesterol concentration, smoking and pacemaker mode (VVIR). /DDDR).

**Conclusions:** Cardiac arrhythmia in patients with permanent pacemakers is always likely to occur, related factors predict 57.8% of the possibility of this event occurring.

## Introduction

One of the main issues in cardiology is heart arrhythmia. In recent years, the treatment of heart arrhythmias has made great progress thanks to advances in technology and a better understanding of the mechanism of arrhythmias ^1^. With more than 60 years of experience, pacemaker treatment for arrhythmias has emerged as a crucial arrhythmia treatment technique in contemporary cardiology. Bradyarrhythmias, tachyarrhythmias, and some other conduction problems are all treated using pacemakers ^2^.

The fundamental objective of pacemaker treatment for bradycardia, which primarily consists of two major syndromes: atrioventricular block and sinus node dysfunction, is to stabilize the heart rate in order to produce good hemodynamics ^3^. Myocardial capture, often referred to as stimulation frequency-dependent myocardial contraction, is a prerequisite for accomplishing that goal. Stimulation must reach a threshold or a specific energy level in order to capture the myocardium. In the electrophysiology of cardiac stimulation, the stimulation threshold, also known as the pacing threshold, is crucial for maintaining stable stimulation, extending pacemaker life, and enhancing patient satisfaction.

Following installation, monitoring the pacemaker becomes a critical matter of care since, apart from ensuring optimal pacemaker operation, aberrant pacemaker disorders must be detected and treated, along with any underlying conditions ^4^. Heart arrhythmia upon implantation is a problem that requires attention in order to guarantee the pacemaker’s maximum efficacy and to identify heart arrhythmia for the purpose of making judgments about more specific therapy, such as atrial fibrillation or both transient and persistent ventricular tachycardia. Since it is challenging to identify arrhythmias following machine deployment through clinical examination, 24-hour ECG monitoring is necessary. However, if the heart arrhythmia is not frequent, it is often not possible to recognize it with just 24 or 48 hours of continuous ECG monitoring. Thanks to advances in science and technology, single-chamber, dual-chamber, and triple-chamber pacemakers and defibrillators all have an important function in helping promptly record significant heart arrhythmias that occur during pacemaker operation ^5^. We can record arrhythmias with these pacemakers in order to verify the diagnosis and choose the best plan of treatment.

When pacemakers are tested today, they are not only checked to make sure they are functioning properly, but they are also checked to see if the patient has any additional arrhythmias that are clinically significant during the pacing process. These tests also check pulse amplitude, pacing frequency, and treatment optimization. Then, decide if treating those arrhythmias with a combination of medications is necessary or if modifying the pacing frequency is sufficient to stop them. This study was conducted to identify some factors predicting arrhythmia after permanent pacemaker placement to treat atrioventricular block and sinus node dysfunction.

## Methods

### Study Participants

This is a cross-sectional descriptive study that involves patients being monitored at Vietnam Thong Nhat Hospital and Thu Duc City Hospital who have permanent pacemakers because of atrioventricular block and sinus node dysfunction. The research project ran from March 2022 to September 2022. Participants who had received an arrhythmia diagnosis in the past were not allowed to participate in the research. The study included 312 research subjects in total who met the disease selection criteria.

### Study Design

Following permanent pacemaker implantation, follow-up visits included clinical and paraclinical evaluations, as well as periodic reexaminations of the study participants. Indicators of age, gender, type of pacemaker, pacemaker brand, diagnosis when placing the pacemaker, and accompanying diseases. Echocardiographic indicators EF (%), LVDd (mm), PAPs (mmHg) were recorded by us through Doppler echocardiographic examination of research subjects. We retrieve pacemaker indicators through the pacemaker’s corresponding software, including parameters on pacing Mode; Pacing rate (Pacing: % of total); Measured Threshold; Sensitivity; Measured Impedance. Corresponding collected laboratory indicators include: Creatinine (mcmol/l)/ GFR; Na (mmol/L); K (mmol/L); Cl (mmol/L); Ca (mmol/L); FT4 (ng/dl); FT3 (ng/ml); TSH (mcUI/ml); Fasting blood glucose (mmol/L); HbA1c (%); Total cholesterol (mmol/L); LDL-c (mmol/L); HDL-c (mmol/L); Triglyceride (mmol/L). We recorded the types of heart arrhythmias in the study subjects during direct examination.

### Statistical Analysis

The distribution of the data was tested using the Skewness test. Logistic regression analysis was used to build a model of the impact of independent factors on arrhythmia events. A p value <0.05 is considered statistically significant. Statistical analyzes were used using SPSS 20.0 software.

## Results

In the study, the ratio of male and female was 56.1% and 43.9%. The average age of the study subjects was 68.8 ± 14.3 years old, of which the age group < 70 years old accounted for the largest proportion, 43.9%; The age group from 70 - 79 years old is 31.4% and the age group older than 79 years old is 24.7%.

Research shows that the main diagnoses for pacemaker placement are sinus node dysfunction (76.3%), second-degree AV block (10.6%), third-degree AV block (13.1%); 62.8% of patients have type 2 diabetes, 52.6% of patients have hypertension, 36.2% of patients have coronary artery disease, 31.1% of patients have dyslipidemia, 20.2% of patients have overweight/obese, 16.7% of patients smoked, 11.5% of patients had chronic kidney disease.

The majority of patients had a dual-chamber pacemaker (68.9%), the most commonly used pacemaker manufacturer was Bio brand (50.8%), followed by Med (27.0%) and St at 22.2%. The modes installed on pacemakers are mainly DDDR (40.7%), DDD (28.5%), VVIR (23.4%), VVI (7.1%) and the remaining mode is AAIR (0.3%) and AAI mode has no patient set.

Research results showed that up to 20.2% of patients had atrial fibrillation, 10.9% of patients had atrial tachycardia, 9.9% of patients had ventricular premature beats, 2.2% of patients had tachycardia. supraventricular, 1.3% of patients had sinus arrest or ventricular tachycardia. The rate of arrhythmias of all types including atrial fibrillation, atrial tachycardia, sinus arrest, ventricular premature beats, ventricular tachycardia and supraventricular tachycardia is 41%. (Table 1) Results in Table 2 show that the variables EF, Potassium, Blood Chlorine, HDL-Cholesterol, smoking, pacemaker mode VVIR, DDDR explain 57.8% of the possibility of heart arrhythmia in patients with permanent pacemakers. Another explanation for the possibility of heart arrhythmia in patients with permanent pacemakers: For every 1 increase in EF, the possibility of heart arrhythmia decreases by 0.97 times; For every 1 increase in potassium, the likelihood of heart arrhythmia increases by 2.18 times; For every 1 increase in blood chloride, the likelihood of heart arrhythmia increases by 1.14 times; For every 1 increase in HDL-Cholesterol, the likelihood of heart arrhythmia increases by 1.65 times; If you smoke, the likelihood of heart arrhythmia increases by 2.34 times; If the pacemaker is in VVIR mode, the possibility of heart arrhythmia is reduced by 0.37 times; If the pacemaker is in DDDR mode, the possibility of heart arrhythmia increases by 2.32 times.

**Table 1.** heart arrhythmia after pacemaker placement.

| Arrhythmia | Yes | No |
| --- | --- | --- |
| Atrial fibrillation | 65 (20.8) | 247 (79.2) |
| Atrial tachycardia | 34 (10.9) | 278 (89.1) |
| Stopped node | 3 (1.0) | 309 (99.0) |
| Ventricular extrasystoles | 31 (9.9) | 281 (90.1) |
| Ventricular tachycardia | 4 (1.3) | 308 (98.7) |
| Supraventricular tachycardia | 7 (2.2) | 305 (97.8) |
| General arrhythmia | 128 (41.0) | 184 (59.0) |

**Table 2.** Logistic regression model of heart arrhythmia events after pacemaker placement.

| Features | $\beta$ | OR<br>(KTC 95%) | p |
| --- | --- | --- | --- |
| EF | -0.03 | 0.97 (0.95 – 0.99) | 0.001 |
| K | 0.78 | 2.18 (1.33 – 3.58) | 0.001 |
| Cl | 0.13 | 1.14 (1.06 – 1.22) | 0.001 |
| HDL-Cholesterol | 0.50 | 1.65 (1.09 – 2.49) | 0.02 |
| Smoking | 0.85 | 2.34 (1.21 – 4.53) | 0.01 |
| VVIR mode pacemaker | -1.00 | 0.37 (0.18 – 0.77) | 0.01 |
| DDDR mode pacemaker | 0.84 | 2.32 (1.33 – 4.05) | 0.001 |

EF: Ejection fraction, K: Potassium, Cl: Blood chloride, HDL-Cholesterol: High Density Lipoprotein- Cholesterol. Predicted to have arrhythmia is 57.8%

## Discussion

Clinical signs of bradycardia can range greatly, from subtle symptoms to obvious syncope. Bradycardia can be divided into two categories: atrioventricular block and sinus node dysfunction. The timing, ventricular rate, general medical state, drugs, and other electrophysiological findings all influence the clinical symptoms. ^6-8^.

In contrast to the findings of Ferreira et al.’s study ^9^, which focused on patients without a history of atrial fibrillation, our investigation revealed a prevalence of atrial fibrillation. The rate of new atrial fibrillation after a pacemaker was implanted was 51.6% (total 345 patients); the findings were comparable to those of Le Tien Dung ^10^, who reported 33.33% of patients with atrial fibrillation; 49.02% of patients had atrial tachycardia; and the rate of ventricular premature beats was comparable to our study. The atrial fibrillation rate found in our study differs from that found in the study by author Cho and colleagues ^11^ who found that atrial fibrillation accounted for 10.3% of all patients who participated. The results of our study, however, are not significantly different from those of the study by author Costa and his colleagues, who found that 47 patients out of 186 patients, or 25.3% of the total, had atrial fibrillation following pacemaker implantation. Additionally, because the authors of this study conducted a multi-year follow-up, they were able to record an additional incidence rate of 5.64 cases per 100 patients per year of follow-up. Atrioventricular block and sinus node dysfunction were the reasons for the subjects’ pacemakers, which may explain the parallels between the study’s results and those of Wu and colleagues ^1^.

Four patients in our study, or 1.3% of the total, experienced sinus arrest during pacemaker checks. The patient’s permanent pacemaker installation is an extremely uncommon occurrence. The medical records of these patients have been removed and re-examined, and four instances of the device running out of battery have been noted. However, either of the Covid-19 pandemic or the distance from home, we have not been to Ho Chi Minh City recently to monitor the gadget on a regular basis. These patients were hospitalized for a new pacemaker because of the concerning circumstances.

Atrial fibrillation is associated with left atrial size and left ventricular systolic function, and its incidence increases progressively with changes in left ventricular anatomical morphology ^12^. Similar to the findings from our investigation of a statistically significant correlation between atrial fibrillation and ejection fraction subgroups, author Qin and colleagues observed a relationship between atrial fibrillation and left ventricular systolic performance with the aforementioned results.

A number of electrolytes are involved in the generation of action potentials across cell membranes; blood potassium levels have a major influence on the variations in action potentials that are most obviously linked to arrhythmias. Our research demonstrated that a one-unit rise in the potassium index, which assesses the risk of arrhythmia, results in a 2.18-fold increase.

The two pacemakers that are most frequently given are VVI and DDD models. These pacemakers offer benefits to patient survival that are comparable. In another study, Healey et al. ^13^, found that patients with a pacemaker set to either AAI or DDD mode had a correlation with a lower risk of atrial fibrillation during the follow-up period. This was especially true for patients diagnosed with sinus node dysfunction In the study by the author Ferreira and colleagues ^9^, new-onset atrial fibrillation was frequently found in patients who had never experienced atrial fibrillation before after the patient had a DDD-type pacemaker installed, and it was linked to long-term death from all causes. This outcome differs from the findings of our research, which revealed a statistically significant correlation between pacemakers of the VVIR or DDDR type and heart arrhythmia in general following pacemaker implantation. This difference may be influenced by the characteristics of the study population, specifically the treatment guidelines and previous reports, where AAI and DDD pacemaker placement is more advantageous and effective. But often due to economic reasons and the circulation of machines in Vietnam, in our study population there were no cases of AAI machines, only one case of AAIR and the vast majority are DDDR two-chamber machines.

### Research limitations

Because of the cross-sectional study design, we were unable to track the long-term results of patients with heart arrhythmias following the implantation of permanent pacemakers.

## Conclusions

The rate of heart arrhythmia after pacemaker placement is 41%, of which 20.2% is atrial fibrillation; 10.9% atrial tachycardia; 1.3% stopped sinus; 9.9% ventricular premature beats; 1.3% ventricular tachycardia and 2.2% supraventricular tachycardia.

Factors that determine the possibility of arrhythmia after permanent pacemaker placement include ejection fraction index, blood potassium, blood chloride, blood HDL-Cholesterol concentration, smoking and pacemaker mode (VVIR/DDDR).

## Data Availability

All data used in this study are available and can be provided to interested parties for non-commercial research purposes, subject to confidentiality agreements and patient privacy regulations. All data have been anonymized to protect patient confidentiality and ensure compliance with medical data protection regulations. The process for requesting and accessing data will adhere to ethical guidelines and applicable laws regarding privacy and data security.

## Ethics Approval and Written Consent from the Patient

This study was approved by the Institutional Review Board of Thu Duc City hospital. The privacy and personal identity information of all participants were protected in accordance with the Declaration of Helsinki. Written informed consent has been obtained from all participants.

## Statement of Informed Consent

The family agreed and consented to the images and medical information being submitted for publication. Written consent was given by the guardians. Institutional approval for the case details was not required, as anonymity was protected.

## References

1. Wu Z, Chen X, Ge J, Su Y. The risk factors of new-onset atrial fibrillation after pacemaker implantation. Herz. 2021;46(Suppl 1):61–68.

2. Alasti M, Machado C, Rangasamy K, et al. Pacemaker-mediated arrhythmias. Journal of arrhythmia. 2018;34(5):485–485.

3. Berruezo A, Mont L, Scalise A, Brugada J. Orthodromic pacemaker-mediated tachycardia in a biventricular system without an atrial electrode. Journal of cardiovascular electrophysiology. 2004;15(9):1100–1100.

4. Allan V, Honarbakhsh S, Casas JP, et al. Are cardiovascular risk factors also associated with the incidence of atrial fibrillation? A systematic review and field synopsis of 23 factors in 32 population-based cohorts of 20 million participants. Thrombosis and haemostasis. 2017;117(5):837–837.

5. Kosmala W, Saito M, Kaye G, et al. Incremental value of left atrial structural and functional characteristics for prediction of atrial fibrillation in patients receiving cardiac pacing. Circulation Cardiovascular imaging. 2015;8(4).

6. Healey JS, Connolly SJ, Gold MR, et al. Subclinical atrial fibrillation and the risk of stroke. The New England journal of medicine. 2012;366(2):120–120.

7. Surawicz B, Childers R, Deal BJ, et al. AHA/ACCF/HRS recommendations for the standardization and interpretation of the electrocardiogram: part III: intraventricular conduction disturbances: a scientific statement from the American Heart Association Electrocardiography and Arrhythmias Committee, Council on Clinical Cardiology; the American College of Cardiology Foundation; and the Heart Rhythm Society. Endorsed by the International Society for Computerized Electrocardiology. Journal of the American College of Cardiology. 2009;53(11):976–976.

8. Tayal B, Riahi S, Sogaard P, et al. Risk of atrial fibrillation after pacemaker implantation: A nationwide Danish registry-based follow-up study. Journal of electrocardiology. 2020;63:153–158.

9. Ferreira V, Portugal G, Viveiros Monteiro A, et al. New onset atrial fibrillation after dual chamber pacemaker implantation: long term predictors. European Heart Journal. 2020;41(Supplement_2).

10. Dũng LT. Nghiên cứu đ ặc điểm rối loạn nhịp tim ở b1EC7nh nhân có hội chứng suy nút xoang trước và sau cấy máy tạo nhịp vĩnh viễn. 2014. Predicted to have arrhythmia is 57.8%

11. Cho MS, Kim J, Kim JH, et al. Clinical, Echocardiographic, and Electrocardiographic Predictors of Persistent Atrial Fibrillation after Dual-Chamber Pacemaker Implantation: An Integrated Scoring Model Approach. PloS one. 2016;11(8):e0160422.

12. Qin D, Heist EK. Atrial fibrillation ablation in congestive heart failure with preserved ejection fraction: Tackling the vicious twins. Journal of cardiovascular electrophysiology. 2020;31(3):689–689.

13. Healey JS, Toff WD, Lamas GA, et al. Cardiovascular outcomes with atrial-based pacing compared with ventricular pacing: meta-analysis of randomized trials, using individual patient data. Circulation. 2006;114(1):11–11.

